# Povidone-iodine ear wash and oral cotrimoxazole for chronic suppurative otitis media in Australian Aboriginal children: a randomised controlled 2x2 factorial design trial

**DOI:** 10.64898/2026.07.20.26358454

**Authors:** Jemima Beissbarth, Christine Wigger, Victor M Oguoma, Amanda J Leach, Ruth Lennox, Sandra Nelson, Hemi Patel, Mark D Chatfield, Kathy Currie, Harvey Coates, Keith Edwards, Heidi C Smith-Vaughan, Kim M Hare, Paul Torzillo, Steven Y.C. Tong, Peter S Morris

**Affiliations:** Menzies School of Health Research, Charles Darwin University; Charles Darwin University - Casuarina Campus: Charles Darwin University; Nil; Northern Territory Department of Health; The University of Queensland; Central Australian Aboriginal Congress; The University of Sydney; Doherty Institute

**Keywords:** Chronic suppurative otitis media, Indigenous, Aboriginal, Cotrimoxazole, Povidone-iodine, Children, Ciprofloxacin, Randomised controlled trial

## Abstract

**Objectives:** To compare the effectiveness of povidone-iodine ear wash compared to no ear wash and oral cotrimoxazole compared to placebo given in addition to standard topical antibiotic treatment (ciprofloxacin drops) for chronic suppurative otitis media (CSOM) in Australian Aboriginal children.

**Methods:** A randomised, parallel, 2 x 2 factorial design, assessor-blinded clinical trial in the remote Northern Territory of Australia. Aboriginal children with confirmed CSOM were eligible to be randomised into four treatment groups, allowing two primary treatment comparisons in a 2-in-1 trial approach. Participants received standard treatment (twice daily cleaning and topical ciprofloxacin drops) plus: i) either 16 weeks of pre-treatment povidone-iodine ear wash or no povidone-iodine ear wash; and ii) either 16 weeks of oral cotrimoxazole or placebo. Central randomisation with allocation concealment and triple-blinding of the oral antibiotic treatment arms was used. The relative risk (RR) and risk difference (RD) were estimated after adjustment for age, community, and the other intervention.

The primary outcome was the proportion of children with any otorrhoea (clinical failure) after 16 weeks of treatment. Secondary outcomes included size of tympanic membrane (TM) perforation and amount of discharge, time to cessation of discharge, proportion of children with respiratory and other pathogens in ear discharge (at baseline and 16 weeks) and hearing levels (at 12 months).

**Findings:** 280 children with CSOM were randomised and 270 had their primary outcome assessed. Clinical failure (presence of any ear discharge) after 16 weeks of treatment was 66/134 (49%) in the povidone-iodine group versus 69/136 (51%) in the no povidone-iodine group (RD= −1% (−12,11), p= 0·93) and 56/134 (42%) in the cotrimoxazole group versus 79/136 (58%) in the placebo group (RD= −16% (−28,-4), p=0·007). The amount of discharge, TM perforation size, the level of hearing impairment, and serious adverse events were not significantly different in both treatment comparisons. Anaerobic growth (24%), *Pseudomonas aeruginosa* (21%) and *Haemophilus influenzae* (17%) were the most common pathogens found in the ear discharge before treatment. Fungi or yeast (24%), *Staphylococcus aureus* (15%), and anaerobic growth (10%) were the common pathogens after 16 weeks of treatment, with no significant differences between groups. At 12 months post-randomisation, 55-60% of children had at least one discharging ear and there was no difference between treatment groups.

**Interpretation:** Povidone-iodine ear washes did not contribute to better ear outcomes in this study. Cotrimoxazole for 16 weeks resulted in more children with clinical improvement to dry ears. Oral cotrimoxazole may play a role in reducing the burden of CSOM in populations with high rates of persistent disease.

**Funding:** This study is funded by the National Health and Medical Research Council (NHMRC) grant number: 1060764.

**Research in context:** *Evidence before this study:* There are three clinical trials relevant to the chronic suppurative otitis media (CSOM) treatments used in this study, two using povidone-iodine, and one of oral cotrimoxazole. Neither of the povidone-iodine trials showed impact from treatment. The trial of 6-12 weeks oral cotrimoxazole demonstrated benefit (15% increased clinical cure). The beneficial effect was not sustained, and the study was not placebo controlled.

*Added value of this study:* This placebo-controlled factorial randomised controlled trial tests the efficacy of oral cotrimoxazole and povidone-iodine in a group of high-risk children. It combines clinical, hearing, and microbiological outcomes. We demonstrate the efficacy of oral cotrimoxazole for up to 16 weeks for children with CSOM.

*Implications of all the available evidence:* The best available evidence supports the use of cotrimoxazole as an adjunct therapy. A course longer than 12 weeks may be required to support clinical cure. The beneficial effect of cotrimoxazole did not persist to the 12 month follow up. The use of topical povidone-iodine as an adjunct therapy for CSOM is not supported.

## BACKGROUND

Chronic suppurative otitis media (CSOM) is the persistent discharge of pus through a perforated tympanic membrane. It remains a significant health issue globally and causes substantial hearing loss and disadvantage that can continue into adulthood.^1,2^ The most recent study (2019) of Australian Aboriginal children in the remote Northern Territory (NT) reports CSOM prevalence of 7%.^3^ Whilst this is a reduction in CSOM compared to a large NT survey in 2001^4^ which reported a prevalence of 15% in children aged 6-30 months, it is well above the 4% threshold classified by the World Health Organization (WHO) as a public health crisis.^5^ Because of the persistent nature of CSOM in our population, treatment is arduous and challenging. In a previous study, 70% of children on current recommended treatment (ciprofloxacin drops after dry mopping the ear canal) did not experience resolution to a dry ear state^6^. These children may continue to have large tympanic membrane perforations throughout their lives. Surgical repair (tympanoplasty) is available in some settings, but access is not universal. While topical antibiotics have been shown to be more effective than oral antibiotics and topical antiseptics, it is unclear whether there are benefits of combining these treatments.^6,7^

A study reporting the use of gently syringing dilute povidone-iodine (0·5%) and dry mopping before applying antibiotic ear drops in Northern Australia described high cure rates.^8^ Pre-treatment povidone-iodine washes were used in both randomised treatment groups. 76% of children in the ciprofloxacin group and 52% in the framycetin (0·5%), gramicidin, dexamethasone (FGD, Sofradex) group had dry ears after 9 days of treatment.^8^ In comparison, a study conducted in a remote NT community school (comparing the same two topical antibiotic drops but without any ear wash) found only 14% in the ciprofloxacin group with dry ears at the 12-28 week follow up.^6^

The only other randomised trial to assess povidone-iodine solution compared a more concentrated povidone-iodine 5% solution (given as topical ear drops) with ciprofloxacin drops. This study of 40 patients with CSOM did not find any difference between these two approaches (with 90% of both groups achieving dry ears by the 16 weeks follow up).^9^

A Dutch study published in 2007 found that the oral antibiotic cotrimoxazole (trimethoprim/sulfamethoxazole) in addition to antibiotic ear drops for 6-12 weeks prolongs ear dryness in CSOM (15% difference (15/47 (23%) vs 23/49 (47%)).^10^ The treatment effect was most pronounced at the 6 week follow up and was reduced if administration of the medication was discontinued. The lack of a placebo medication was a potential source of bias.

The aim of this randomised clinical trial was to determine whether, amongst Australian Aboriginal children with CSOM, either povidone-iodine ear wash (compared to no povidone-iodine ear wash) or oral cotrimoxazole (compared to placebo) given in addition to standard topical antibiotic treatment reduces the proportion of children with ear discharge after 16 weeks of treatment. We used a 2×2 factorial design so that we could assess the effect of both interventions at the same time (a 2-in-1 trial approach). We assumed that there would be no interaction between the two interventions being assessed.

## METHODS

### Study design

The study design and rationale for this study have been previously published.^11^ In summary, we conducted a 2 x 2 factorial design, randomised, assessor-blinded, controlled clinical trial across two urban and 26 remote communities in the NT between 2015 and 2018. The phone-activated 1:1:1:1 computer-generated randomisation process was centralised and provided by the NHMRC Clinical Trials Centre, Sydney (who were not directly involved with the trial). The random allocation sequence was stratified by community and age (<4 years; 4-10 years; and >10 years old) and used a minimisation algorithm. Allocation was concealed throughout the trial. We compared current standard treatment of CSOM (twice daily dry mopping, ciprofloxacin drops and weekly clinic review) with two adjunctive therapies; pre-treatment povidone-iodine ear wash (or no povidone-iodine ear wash) and/or oral cotrimoxazole (or placebo). Standard treatment, povidone-iodine ear wash, and cotrimoxazole or placebo, were all administered for 16 weeks. It was not possible to blind the topical treatment. Allocation of the oral intervention was blinded from participants, investigators, and those providing routine clinical care by using a placebo with a non-active formulation of the exact colour, taste and appearance as the intervention.

### Participants and setting

Children aged 2 months to 17 years of age living in remote or urban NT communities with a diagnosis of CSOM (middle ear discharge present for longer than 2 weeks with perforation of the tympanic membrane of >2%; presence of tympanic membrane perforation and discharge confirmed by research team) in at least one ear were eligible. Data collection occurred between 2015 and 2018. Children were excluded for the following: i) previously randomised in this study; ii) ciprofloxacin, cotrimoxazole or iodine allergy; iii) mastoid or tympanoplasty surgery in the preceding 12 months; iv) ear surgery scheduled in the next four months; v) congenital ear or hearing problems; vi) known immunodeficiency; or vii) pregnant.

### Ethics and consent

This study was approved by the Human Research Ethics Committee of the Northern Territory Department of Health and the Menzies School of Health Research (HREC-2014-2170), and the Central Australian Human Research Ethics Committee (CAHREC-14-228). Each participating community council/board provided written approval of the study. Parent/guardian consent (and assent from children over 10 years of age) was obtained prior to enrolment as well as ongoing verbal confirmation during the study participation period.

### Intervention

Children with confirmed CSOM were randomised to receive current standard treatment of CSOM (twice daily dry mopping followed by insertion of ciprofloxacin drops and weekly clinic review) plus one of two interventions:

1: Topical aqueous povidone-iodine 10%, diluted in clean, room temperature water (1:20). Each discharging ear was syringed twice daily using a 20ml syringe for up to 16 weeks or until the ear was dry for 14 days (confirmed on two clinical examinations one week apart by study or clinic staff).
2: Cotrimoxazole suspension (or tablet for children over 12 years of age) or placebo. Cotrimoxazole suspension was given as 4mg/kg per dose (of trimethoprim component) twice a day orally for 16 weeks (or if the treating clinician or research team decided that there was no ear discharge for >3 days as per the Otitis Media Guidelines for Australian Aboriginal and Torres Strait Islander children^12^).

The placebo was administered in an identical manner. Treatment was recommenced if discharge became present again.

Other ear conditions (such as AOM) were managed according to best practice treatment guidelines. Intercurrent illnesses were referred to the participant’s local clinic for management.

### Primary outcome measures

The primary outcome measure was the proportion of children with clinical failure after 16 weeks of treatment. Clinical failure was determined by an ear examination and defined as the presence of any ear discharge (otorrhea) in the ear canal at 16 weeks.

### Secondary outcome measures

In this paper, we report the following prespecified secondary outcome measures at 16 weeks: i) failure to improve (determined presence of discharge); ii) change in perforation size (determined by comparison with standardised assessment at baseline); iii) adherence to recommended management using pharmacy dispensing records and participant reported use (diary); iv) presence of recognised bacterial pathogens in ear discharge (determined by standard microbiological culture and antimicrobial sensitivity methods); v) complications (defined as development of a related illness requiring additional medical treatment); vi) side effects (defined as development of illness requiring cessation of prescribed treatments): and vii) audiology levels measured at 12 months (plus or minus 6 months) and summarised as the pure tone 4 frequency average hearing level at 500, 1000, 2000 and 4000 Hz in the worse hearing ear. If fewer frequencies were tested, the average of those tested were used (ie; 1, 2, 3 or 4 frequencies). Hearing data was collected via medical file review. Audiometric assessments were performed by government audiologists.

Secondary microbiological outcomes (carriage of vaccine serotype capsular pneumococci, non-vaccine serotype capsular pneumococci) not reported here will be described in a more detailed separate publication of the microbiologic assessments (including antibiotic susceptibility testing of otopathogens).

### Clinical assessment

All recruitment, treatment, and clinical assessment was conducted by trained ear research nurses. After the eligibility assessment and consent had been completed, the research nurse contacted the NHMRC Clinical Trials Centre by phone to receive the randomised allocation to treatment. Ear examinations were made at baseline, 16 weeks and 12 months follow-up. Otoscopic and tympanometry findings were recorded on standardised forms and all video recordings of ear state were saved for review by a senior ear researcher as required. Assessments were made using Voroscope and Siegle speculum pneumatic otoscopy (Vorotek, VIC, AUS), Welch Allyn (Skaneateles, NY, USA) and MedRx (Largo, FL, USA) video-otoscopy and GSI 38/39 tympanometry (Grason-Stadler, Eden Prairie, Minn, USA).

### Microbiology

Ear swabs (for all discharging perforations) were collected at baseline, 16 weeks (canal swab collected if no discharge), and 12 months (canal swab collected if no discharge) follow-up examinations. Non-flocked rayon swabs (Copan Italia, Italy) were used. Post collection swabs were placed into vials containing 1ml of skim milk, tryptone, glucose, glycerol broth (STGGB), placed immediately into a liquid nitrogen dry shipper, transported frozen to the laboratory and transferred into a −80°C freezer. Culture methods were designed for the detection of otopathogens (*Streptococcus pneumoniae,* non-typeable *H. influenzae* (NTHi) and *Moraxella catarrhalis*) and other target species *(Pseudomonas aeruginosa, S. pyogenes, S. aureus, Proteus species*, fungi and yeasts, coliforms, and general anaerobes). Aliquots (10µL) were inoculated onto chocolate agar, horse blood agar, colistin nalidixic acid agar (CNA) and MacConkey biplates, and bacitracin vancomycin, clindamycin, chocolate agar (BVCCA) plates incubated at 37°C in 5% CO2 for 18-24 hours, pseudomonas agar incubated at 37°C in air and Sabouraud-CG slope at 28°C in air for 6 weeks, observed weekly. An anaerobe plate was inoculated with a metronidazole disc (5ug) at the first set of streaks from the primary zone and incubated at 37°C for 48 hours with an anaerobe pack. For all bacteria of interest, one random colony was selected from the dominant morphological type and one other colony of any different morphological type if present. All bacteria of interest were identified using standard methods (Supplement 1).

### Statistical analysis

Reporting follows the analysis plan in the protocol publication.^11^ The factorial analysis used a pre-specified 2-in1 trial approach (at-the-margins) that assumed no interaction effect. The objective was to evaluate the main effect of each intervention (povidone-iodine wash and oral antibiotic cotrimoxazole) as separate adjunctive treatments to standard care alone ie. adjunctive povidone-iodine wash vs. no adjunctive treatment and cotrimoxazole vs. placebo. The estimand of interest for each intervention (the target intervention effect) was the effect in the absence of the other factor. Our planned sample size was 280 participants (140 patients per treatment group, 70 per arm). The study was powered at 90% to detect improved outcome in an additional 20% of children for each intervention (the two main effect comparisons) allowing for a 10% loss to follow-up. We assumed that 70% of children would fail to respond without adjunctive treatment.^6,13^ All participants randomly assigned to receive interventions were considered to constitute the intention-to-treat (ITT) population for the primary and secondary analyses. Our primary outcome assessment used complete case analysis and did not impute any missing data. We also described an alternative ITT analysis that assumed any children lost to follow up were clinical failures. For the primary and secondary dichotomous outcomes, binomial regression models with logarithmic and identity links were used to estimate the main effects of the two interventions. We prespecified that the analysis would assume that there was no interaction between the two main effects. The relative risk and the risk difference with the 95% confidence interval (CI) are reported for the crude and adjusted estimates. Adjustment for community and age (stratification variables), and the other intervention (as a covariate) was pre-specified. The possibility of an interaction effect between the two interventions (povidone-iodine wash and cotrimoxazole) was assessed by adding an interaction term to the log-binomial regression model. For hearing levels, the mean difference between groups and 95% CI are reported for the worse ear. We estimated differences in the primary outcome across specified subgroups by adding appropriate interaction terms to the regression models. Two-sided values of p<0·05 were considered significant. We pre-specified that there would be no interim analyses of the intervention main effects unless requested by the Data Safety Monitoring Committee. The committee met at 6-12 month intervals throughout the trial. All analysis was performed using Stata 15.1 (StataCorp, College Station, TX)

## RESULTS

1732 assessments of high-risk children were undertaken in 26 remote communities and two urban centres. Of these 1732, 355 child assessments in 24 communities (20%) were discharging perforations. 75 eligible assessments did not result in randomisation as a parent or child declined to participate or met one of the exclusion criteria (Figure 1). 280 (16%) children were randomly allocated to a treatment group. 279 were used in the analysis (one child was excluded post-randomisation for incorrect diagnosis of CSOM on review of the otoscopy video). We collected clinical outcome data (ear examination) on 270/279 (97%) of participants at 16 weeks and 278/279 (99%) at 12 months (Fig 1). Loss to follow-up was due to participants not being in their respective community at the time of our nurse visits (up to three follow up attempts were made, depending on the accessibility of the community).

**Figure 1:**
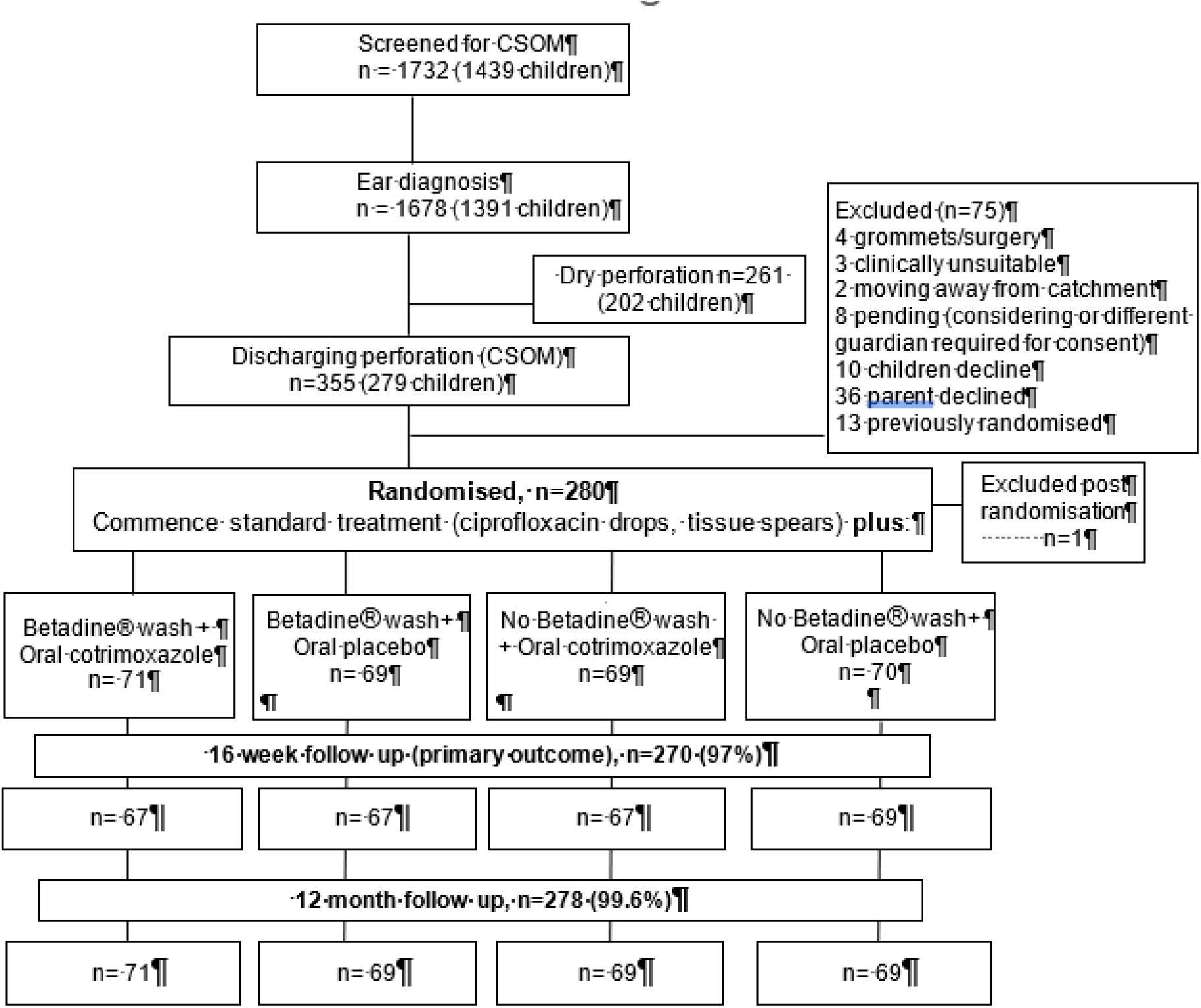
Randomisation and allocation

**Figure 1:**
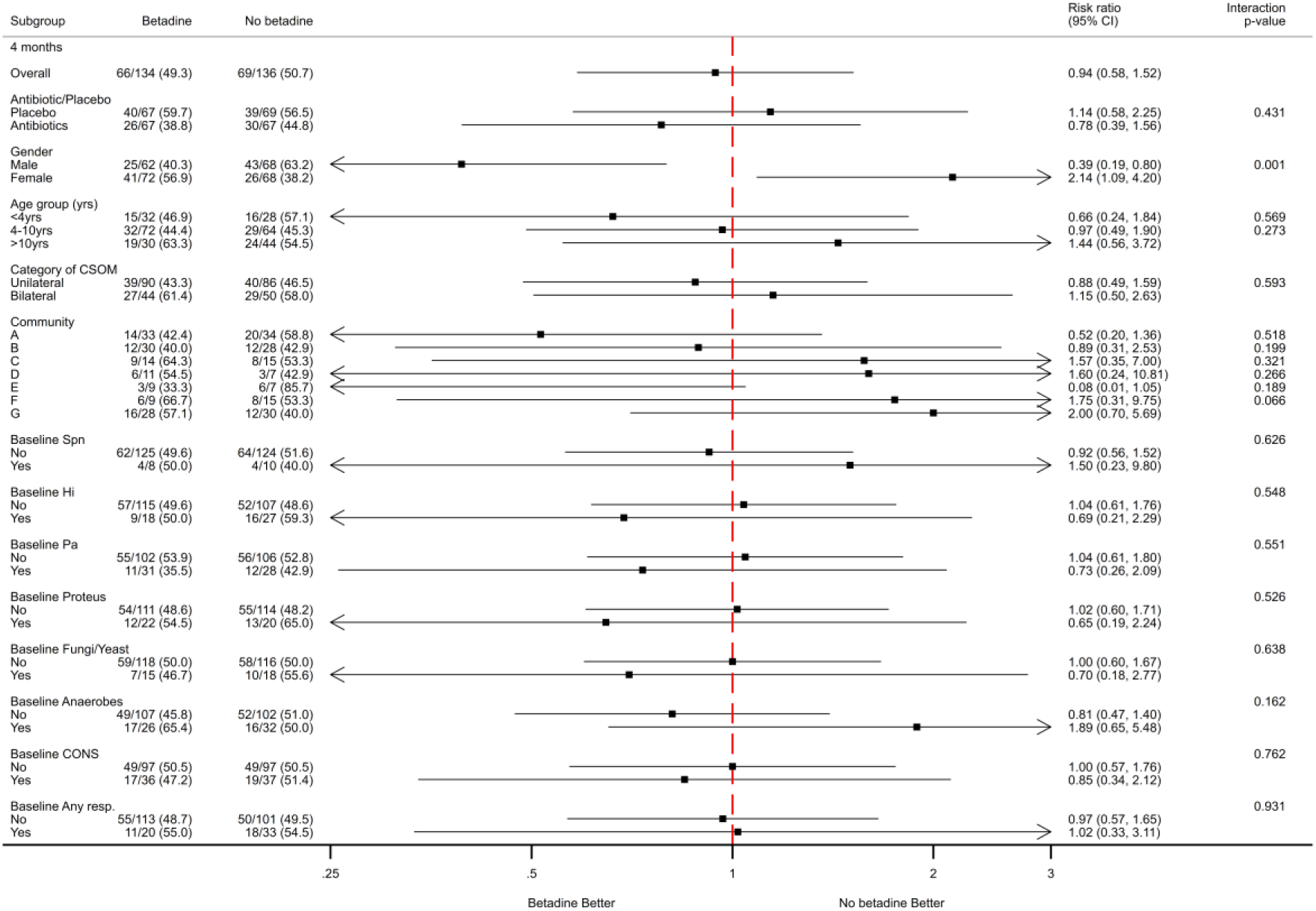
16 week outcomes for povidone-iodine vs no povidone-iodine

### Baseline characteristics

Of the 279 children, 135/279 (48%) were male. Age ranged from 6 months to 18 years (mean age 7 years). 274/279 (98%) of the children in this study lived in a remote Aboriginal community in the Northern Territory (most requiring off road vehicles or planes to reach). English was not usually their first language. Maternal smoking was reported in 154/279 (55%) of respondents. 83/233 (36%) of mothers reported that they had experienced “runny ears” as a child, 98/251 (39%) of study children had a sibling who had runny ears and 94/229 (41%) of children had an immediate family member who had runny ears. 78/261 (30%) of respondents (parent or guardian) were in paid employment and 200/257 (78%) were receiving income support. 215/266 (81%) of children had been (or still were) breast fed. Three children (1%) had a (unilateral) discharging myringotomy tube (grommet) in situ at the time of their enrolment.

Baseline characteristics are reported in Table 1, according to the four randomly allocated treatment groups: i) povidone-iodine wash plus oral cotrimoxazole; ii) povidone-iodine wash plus oral placebo; iii) no povidone-iodine wash plus oral cotrimoxazole; and iv) no povidone-iodine wash and oral placebo. Where whole of life data were available (N=182), mean age of first perforation was 0.9 years (range 0.07-6.1 years).

**Table 1:** Baseline characteristics.

|  |  | No povidone-iodine/<br>placebo<br>N=70 | No Povidone-iodine/<br>Cotrimoxazole<br>N=69 | Povidone-iodine/<br>placebo<br>N=69 | Povidone-iodine/<br>Cotrimoxazole<br>N=71 |
| --- | --- | --- | --- | --- | --- |
| Gender | Male | 32/70 (46%) | 37/69 (54%) | 28/69 (41%) | 38/71 (54%) |
|  | Female | 38/70 (54%) | 32/69 (46%) | 41/69 (59%) | 33/71 (46%) |
| Age (mean, years (SD)) |  | 8 (4) | 7 (4) | 7 (4) | 7 (4) |
| Age (year group) | <4yrs | 13/70 (19%) | 17/69 (25%) | 21/69 (30%) | 19/71 (27%) |
|  | 4-10yrs | 35/70 (50%) | 34/69 (49%) | 31/69 (45%) | 38/71 (54%) |
|  | >10yrs | 22/70 (31%) | 18/69 (26%) | 17/69 (25%) | 14/71 (20%) |
| Weight (kg) (SD) |  | 25 (13) | 24 (15) | 24 (13) | 22 (12) |
| CSOM category | Unilateral | 48/70 (69%) | 41/69 (59%) | 46/69 (67%) | 49/71 (69%) |
|  | Bilateral | 22/70 (31%) | 28/69 (41%) | 23/69 (33%) | 22/71 (31%) |
| Worst perforation | 3-10% | 16/70 (23%) | 19/69 (28%) | 16/69 (23%) | 24/71 (34%) |
|  | 11-25% | 28/70 (40%) | 20/69 (29%) | 23/69 (33%) | 24/71 (34%) |
|  | 26-50% | 14/70 (20%) | 20/69 (29%) | 22/69 (32%) | 14/71 (20%) |
|  | 51-75% | 8/70 (11%) | 7/69 (10%) | 5/69 (7%) | 6/71 (8%) |
|  | >75% | 4/70 (6%) | 3/69 (4%) | 3/69 (4%) | 3/71 (4%) |
| Median worst perforation size % (IQR) |  | 21 (12-38) | 22 (9-38) | 25 (12-35) | 20 (8-30) |
| Hearing (worst) |  | n=44 | n=45 | n=39 | n=40 |
|  | normal | 4/44 (9%) | 4/45 (9%) | 6/39 (15%) | 6/40 (15%) |
|  | mild | 14/44 (32%) | 14/45 (31%) | 11/39 (28%) | 15/40 (38%) |
|  | moderate | 26/44 (59%) | 27/45 (60%) | 21/39 (54%) | 18/40 (45%) |
|  | severe | 0/44 (0%) | 0/45 (0%) | 1/39 (3%) | 1/40 (3%) |
| Mean worst hearing loss dB (SD) |  | 34 (10) | 34 (10) | 32 (11) | 32 (11) |
| Ear discharge |  | n=68 | n=69 | n=69 | n=70 |
|  | *P. aeruginosa* | 16/68 (24%) | 12/69 (17%) | 19/69 (28%) | 12/70 (17%) |
|  | NTHi | 13/68 (19%) | 14/69 (20%) | 12/69 (17%) | 7/70 (10%) |
|  | Swarming proteus | 10/68 (15%) | 10/69 (14%) | 15/69 (22%) | 9/70 (13%) |
|  | *S. aureus* | 12/68 (18%) | 15/69 (22%) | 7/69 (10%) | 5/70 (7%) |
|  | *S. pneumoniae* | 4/68 (6%) | 6/69 (9%) | 4/69 (6%) | 4/70 (6%) |
|  | Anaerobic growth | 12/68 (18%) | 22/69 (32%) | 11/69 (16%) | 19/70 (27%) |
|  | Fungi or yeast | 8/68 (12%) | 10/69 (14%) | 8/69 (12%) | 7/70 (10%) |
|  | Coag-neg Staph | 21/68 (31%) | 16/69 (23%) | 16/69 (23%) | 22/70 (31%) |
|  | Any growth* | 68/68 (100%) | 69/69 (100%) | 66/69 (96%) | 66/70 (94%) |

\*Any growth is defined as any cultured bacteria or fungi/yeasts;

At time of enrolment, parents reported 22% of children had experienced discharge from their perforation for >12 months, 14% for >6 months, 29% for >3months and 20% for >1 month, 14% were unsure. Also by parent report, 25% of children were being treated for ear problems, 20% were using drops, 14% tissue spears, 8% oral medications, 1% bush medicines, <1% betadine wash, and <1% an injected treatment. By clinic record review, 17% of children were on recommended ear treatment (topical ciprofloxacin or equivalent) at the time of enrolment into the study.

At baseline 184/279 (66%) of children had unilateral perforations and 170/279 (61%) had perforations smaller than 25% of the tympanic membrane. 168/279 (60%) of children had a hearing test available (up to 2 years prior and 1 month after randomisation). Hearing was normal (0-20 dB) in 5/168 (12%) of the children, while 69/168 (32%) had mild (21-30 dB), 92/168 (55%) had moderate (31-60dB) and 2/168 (1%) had severe (61-90dB) hearing impairment. The mean hearing loss was above 30dB, which WHO states is a disabling hearing loss for children. Across all groups, age, gender and weight were balanced and severity of ear disease and hearing impairment. Ear microbiological culture results were also similar. Anaerobic growth (23%), *P. aeruginosa* (21%) and NTHi (17%) were the most common pathogens found in the ear discharge (by child) before study treatment commenced. *Proteus* species (16%), *S. aureus* (14%), fungi and yeasts (12%) and pneumococci (7%) were also cultured.

At baseline, 19/279 (7%) children had chronic conditions requiring regular antibiotic treatment. Most of these children had chronic suppurative lung disease (bronchiectasis) or rheumatic heart disease / acute rheumatic fever.

### Clinical outcomes

Primary outcome was clinical failure (presence of any ear discharge) after 16 weeks of treatment. In the povidone-iodine group 66/134 (49%) children were clinical failures versus 69/136 (51%) in the no povidone-iodine group (RD= −0·5% (−12,11), p= 0.88) (table 2). We found that with twice daily cotrimoxazole for up to 16 weeks, dry ears were observed in 56/134 (42%) children in the cotrimoxazole group versus 79/136 (58%) in the placebo group (RD=-16% (−28,-4), p=0·015) (Table 3). There was no statistically significant interaction between povidone-iodine and cotrimoxazole on the primary outcome. The interaction RR (ratio of relative risks) at 16 weeks was 0.82 (0.5, 1.34, p=0·43).

**Table 2.** Primary outcome (clinical failure): Povidone-iodine versus no povidone-iodine ear wash at 16 weeks.

|  | Povidone-iodine<br>(n=134) | No Povidone-iodine (n=136) | Risk Ratio<br>(95% CI) | Risk<br>Difference<br>(95% CI) | p-value |
| --- | --- | --- | --- | --- | --- |
| Crude | 66 (49%) | 69 (51%) | 0.97 (0.76,1.23) | -1.5% (-10,10) | 0.81 |
| Adjusted# |  |  | 1.02 (0.80,1.31) | -0.5% (-12,11) | 0.88 |
| ITT^ | 72/140 (51%) | 72/139 (52%) | 1.00 (0.82,1.22) | -1.8% (-10,10) | 0.95 |

#Adjusted for age, community, and the other intervention (cotrimoxazole)
^ ITT – children not examined assumed to be clinical failures

**Table 3.**
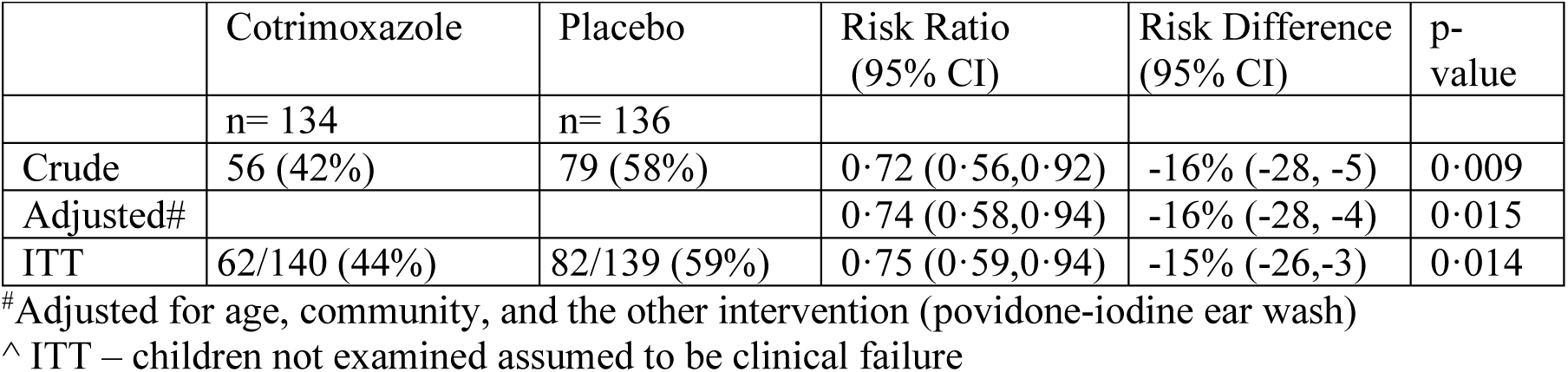
Primary outcome (clinical failure): Cotrimoxazole versus placebo at 16 weeks.

**Table 4.** Clinical and microbiology outcomes at 16 weeks, by povidone-iodine randomisation.

|  |  | Povidone-iodine<br>n=134 | No Povidone-iodine<br>n=136 | p-value |
| --- | --- | --- | --- | --- |
| Worst ear | Normal | 2/134 ( 1%) | 4/136 ( 3%) | 0.63 |
|  | OME | 13/134 (10%) | 7/136 ( 5%) |  |
|  | AOMwiP | 2/134 ( 1%) | 2/136 ( 1%) |  |
|  | Dry perf | 53/134 (40%) | 56/136 (41%) |  |
|  | CSOM | 64/134 (48%) | 67/136 (49%) |  |
| CSOM | Unilateral | 40/64 (63%) | 48/67 (72%) | 0.27 |
|  | Bilateral | 24/64 (38%) | 19/67 (28%) |  |
| Worst perforation | Intact | 18/132 (14%) | 13/135 (10%) | 0.45 |
|  | <=2% | 3/132 ( 2%) | 3/135 ( 2%) |  |
|  | 3-10% | 36/132 (27%) | 27/135 (20%) |  |
|  | 11-25% | 31/132 (23%) | 38/135 (28%) |  |
|  | 26-50% | 25/132 (19%) | 37/135 (27%) |  |
|  | 51-75% | 15/132 (11%) | 15/135 (11%) |  |
|  | >75% | 4/132 ( 3%) | 2/135 ( 1%) |  |
| Ear discharge |  | n=134 | n=131 |  |
|  | *P. aeruginosa* | 10/134 ( 7%) | 6/131 ( 5%) | 0.32 |
|  | NTHi | 15/134 (11%) | 8/131 ( 6%) | 0.14 |
|  | *Proteus* | 8/134 ( 6%) | 10/131 ( 8%) | 0.59 |
|  | *S. aureus* | 18/134 (13%) | 21/131 (16%) | 0.55 |
|  | Coag-neg Staph | 51/134 (38%) | 38/131 (29%) | 0.12 |
|  | *S. pneumoniae* | 6/134 ( 4%) | 4/131 ( 3%) | 0.54 |
|  | Anaerobic growth | 15/134 (11%) | 13/131 (10%) | 0.74 |
|  | Fungi or yeast | 34/134 (25%) | 31/131 (24%) | 0.75 |
|  | Any growth* | 127/134 (95%) | 121/131 (92%) | 0.42 |

\*Any growth is defined as any cultured bacteria or fungi/yeasts

### Secondary outcomes (perforation size and discharge volume at 16 weeks)

Across all groups combined, 50% of children had dry ears in the study after 16 weeks. At 16 weeks, there was a trend for mean perforation size by worst ear, to reduce in all groups, with 12% tympanic membrane healed and 2% had a perforation size of less than 2%. The mean number of perforations ≥50% of the tympanic membrane increased from 9% to 11% from baseline to follow up. The discharge compared with baseline was able to be assessed in 267 children (nine children lost to follow up, and three where data were not available at either baseline or follow up for assessment). In the povidone iodine group, 87/131 children had less discharge at follow up (RD=-6% (−17, 5; p=0·257), and in the cotrimoxazole group 97/132 had less discharge at follow up compared to baseline (RD=6 (−3,19; p=0·179) (Figures 2, 3). Possible subgroup effects were described, and we did not find any subgroup effects that would change practice (16 weeks Figures 2 and 3; 12 months Figures 4 and 5)

**Figure 2:**
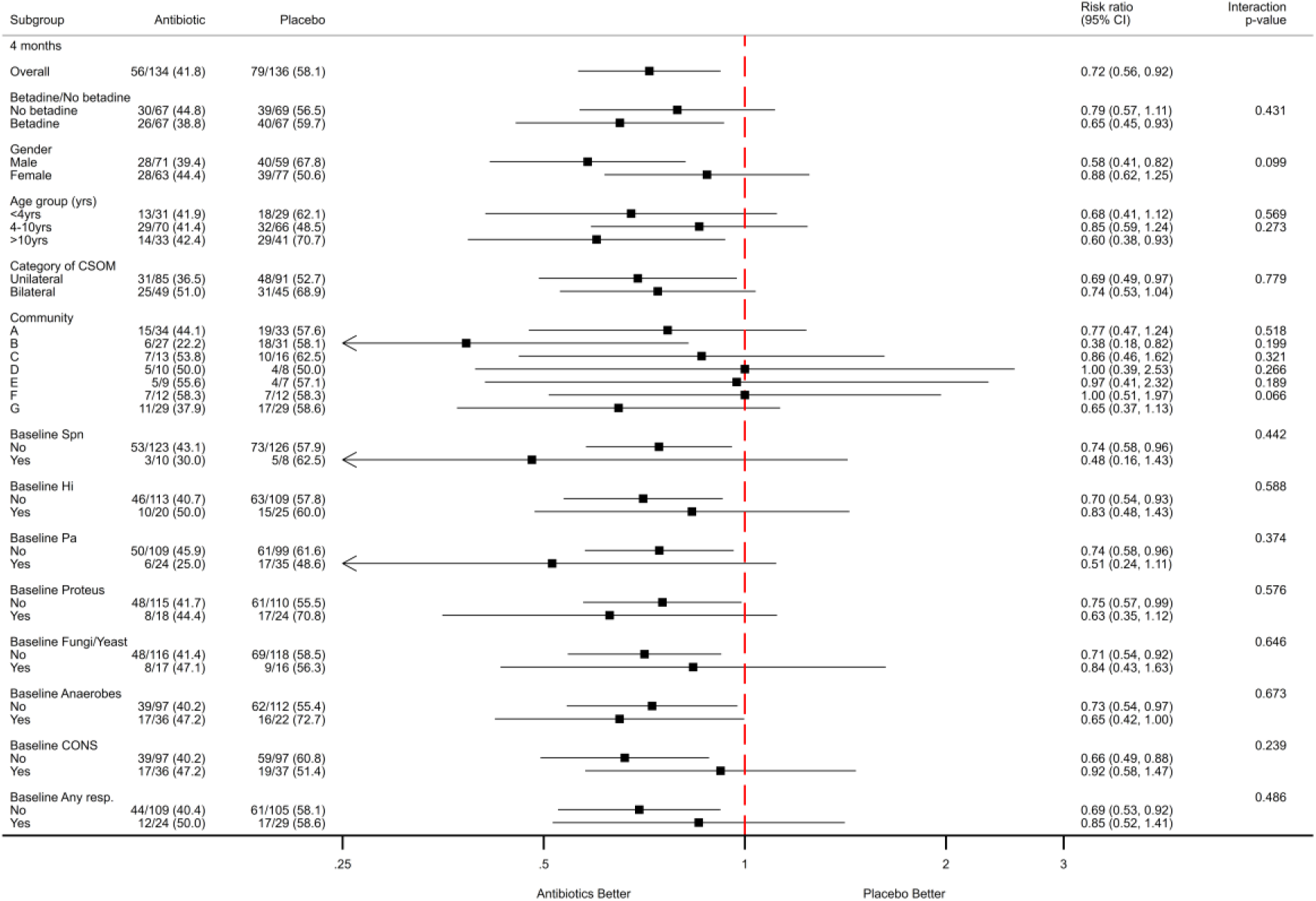
16 week outcomes for cotrimoxazole vs placebo.

**Figure 3:**
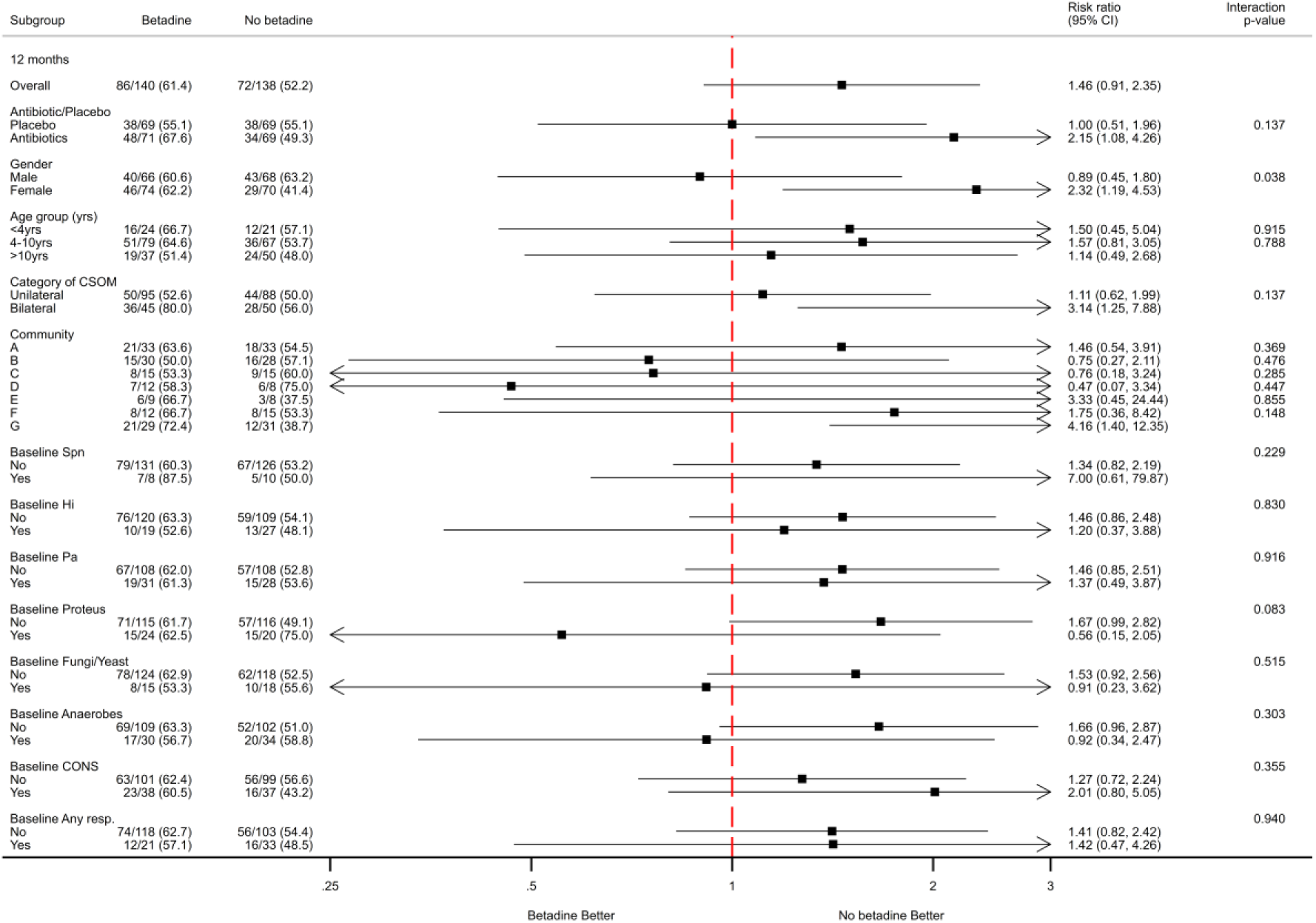
12 month outcomes for povidone-iodine vs no povidone-iodine.

**Figure 4:**
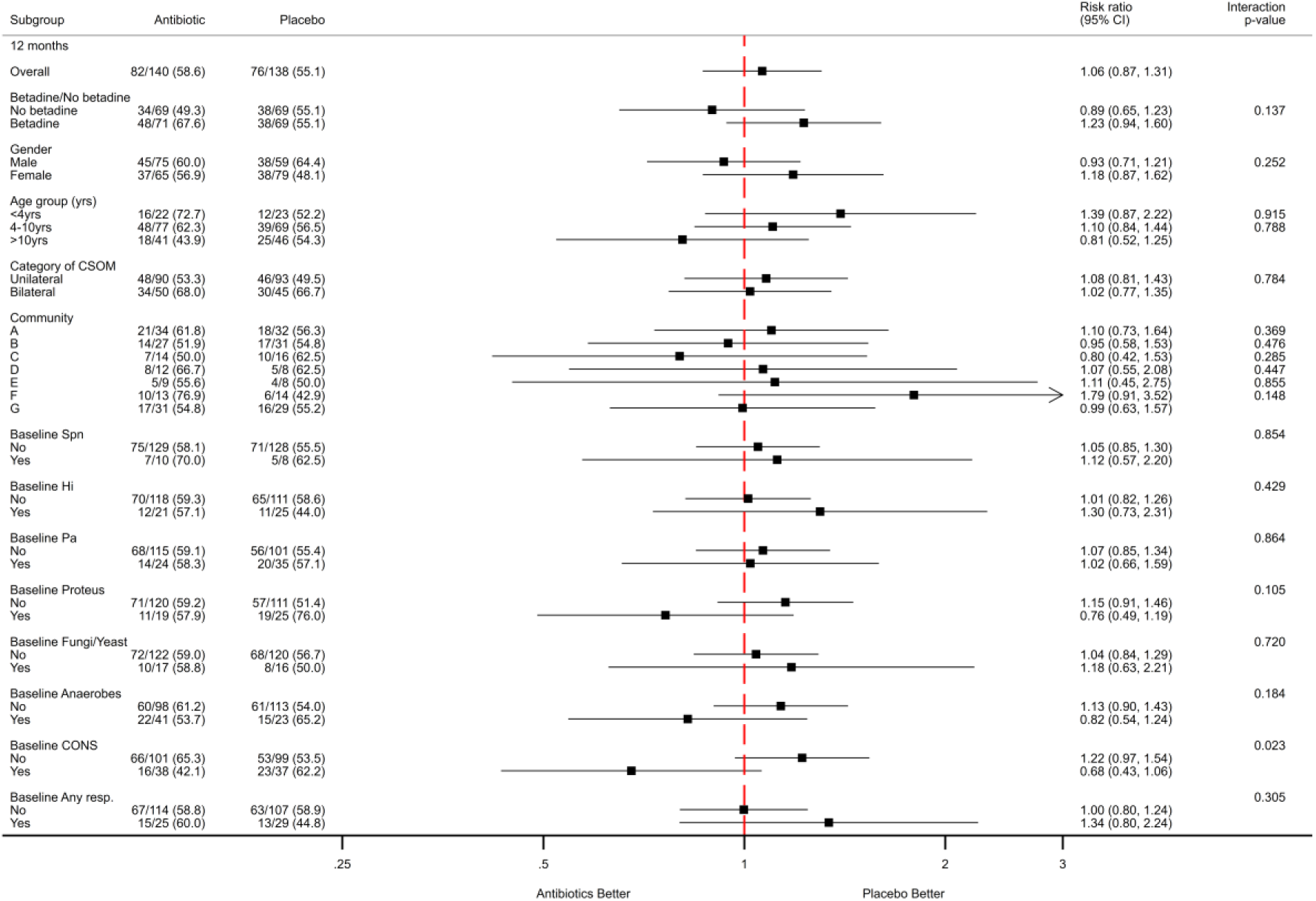
12 month outcomes for cotrimoxazole vs placebo.

At baseline, 184 (66%) children had unilateral CSOM. 7 (3%) of 278 children had developed CSOM in the opposite ear at the end of treatment (from OME or normal), and 18 (6%) children with a unilateral dry perforation at baseline were discharging from that ear at the end of treatment. For children with dry ears at 16 weeks it was not possible to estimate how long the ears had been dry.

### Clinical outcomes at 12 months

At the 12 month follow up, 158 (57%) of 278 children who were assessed had discharging ears. This was not significantly different by randomisation group [povidone-iodine vs no povidone-iodine: 86/140 (61%) vs 72/138 (52%); Adjusted RD 8% (−3, 20); p= 0·163. Cotrimoxazole vs placebo 82/140 (59%) vs 76/138 (56%); Adjusted RD 3% (−8, 15); p=0·570. The interaction RR (ratio of relative risks) at 12 months was 1.37 (0.9, 2.08, p=0·14) (Figures 4, 5; supplement 1, table 3).

### Adherence to recommended management

Paper-based parent dosing records were initiated at the beginning of the trial. However, most parents were not able to return the record at follow up visits (or at end of treatment), and this was abandoned. Medication dispensing records indicate that 36% of children had documentation of dispensing of >75% of medication required to complete the 16 week course of antibiotic or placebo, 21% had documentation of 50-74%; 24% had documentation of 25-49%; and 19% had documentation of <25%.

### Other bacterial infections

98/279 (35%) children had a total of 158 prescribed additional antibiotic treatments within the intervention period of 16 weeks. 11% of antibiotic prescriptions were for respiratory illness (including tonsillitis and pneumonia), 30% for other ear conditions such as AOM, 43% for bacterial skin infections (including dog bites, infected scabies and impetigo), and 14% for other conditions including trachoma prevention, dental related infections, unspecified febrile illnesses, diarrhoeal illnesses, and urinary tract infections.

### Serious adverse events

There were 69 hospitalisations during the study. This included any ENT surgery, any hospital admission longer than 24 hours, and medical evacuations (to central hospitals) from remote primary healthcare facilities for treatment (Table 7, supplement 1, table 5). Of the hospital admissions, 46% were for ear conditions or surgeries (AOM, CSOM, pain, myringotomy, grommet insertion, myringoplasty, cholesteatoma, polyp removal), 14% for respiratory conditions (pneumonia, respiratory infections, asthma), 10% for skin and soft tissue infections, 7% for other surgeries, 4% for unspecified fevers, 3% for gastrointestinal illness, 4% for injury, and 10% for other conditions. None of these hospitalisations were thought to be directly related to either of the interventions.

**Table 5.** Clinical outcomes in at 16 weeks, by cotrimoxazole randomisation.

|  |  | Cotrimoxazole | Placebo | p-value |
| --- | --- | --- | --- | --- |
|  |  | n=134 | n=136 |  |
| Worst ear | Normal | 4/134 ( 3%) | 2/136 ( 1%) | 0·080 |
|  | OME | 10/134 ( 7%) | 10/136 ( 7%) |  |
|  | AOMwiP | 1/134 ( 1%) | 3/136 ( 2%) |  |
|  | Dry perf | 64/134 (48%) | 45/136 (33%) |  |
|  | CSOM | 55/134 (41%) | 76/136 (56%) |  |
| CSOM | Unilateral | 36/55 (65%) | 52/76 (68%) | 0·72 |
|  | Bilateral | 19/55 (35%) | 24/76 (32%) |  |
| Worst perforation | Intact | 15/132 (11%) | 16/135 (12%) | 0·76 |
|  | <=2% | 2/132 ( 2%) | 4/135 ( 3%) |  |
|  | 3-10% | 35/132 (27%) | 28/135 (21%) |  |
|  | 11-25% | 34/132 (26%) | 35/135 (26%) |  |
|  | 26-50% | 32/132 (24%) | 30/135 (22%) |  |
|  | 51-75% | 12/132 ( 9%) | 18/135 (13%) |  |
|  | >75% | 2/132 ( 2%) | 4/135 ( 3%) |  |
| Ear discharge | Collected | n= 129 | n= 136 |  |
|  | *P. aeruginosa* | 9/129 ( 7%) | 7/136 ( 5%) | 0·53 |
|  | *NTHi* | 12/129 ( 9%) | 11/136 ( 8%) | 0·73 |
|  | *Proteus* | 8/129 ( 6%) | 10/136 ( 7%) | 0·71 |
|  | *S. aureus* | 19/129 (15%) | 20/136 (15%) | 1·00 |
|  | Coag-neg Staph | 48/129 (37%) | 41/136 (30%) | 0·22 |
|  | *S. pneumoniae* | 7/129 ( 5%) | 3/136 ( 2%) | 0·17 |
|  | Anaerobic growth | 13/129 (10%) | 15/136 (11%) | 0·80 |
|  | Fungi or yeast | 36/129 (28%) | 29/136 (21%) | 0·21 |
|  | Any growth* | 120/129 (93%) | 128/136 (94%) | 0·72 |

\*Any growth is defined as any cultured bacteria or fungi/yeasts

**Table 6.** Additional antibiotics in the intervention period, by cotrimoxazole randomisation.

| Condition treated | Placebo<br>N=136 |  | Cotrimoxazole<br>N=134 |  | Absolute<br>difference<br>% (95% CI) | P-value |
| --- | --- | --- | --- | --- | --- | --- |
|  | n (%) | 95% CI | n (%) | 95% CI |  |  |
| Total additional treatments | 57 (42) | (34, 51) | 41 (30) | (23, 39) | -11 (-23, 0) | 0·058 |
| Ears | 17 (13) | (8, 19) | 12 (9) | (5, 15) | -4 (-11, 4) | 0·433 |
| Skin / soft tissue infections | 24 (18) | (12, 25) | 19 (14) | (9, 21) | -4 (-12, 5) | 0·507 |
| Respiratory | 7 (5) | (2, 10) | 5 (4) | (1, 9) | -1 (-6, 4) | 0·769 |
| ARF / RHD | 1 (1) | (0, 4) | 0 (0·0) | (0, 3) | -1 (-2, 1) | 1·000 |
| Other | 8 (6) | (3, 11) | 6 (5) | (2, 10) | -1 (-7, 4) | 0·785 |

**Table 7.** Serious Adverse Events summary by randomisation group.

| 4 months | Placebo | Cotrimoxazole | No Povidone-iodine | Povidone-iodine |
| --- | --- | --- | --- | --- |
|  | N=136 | N=134 | N=136 | N=134 |
| ENT | 8/136 (6%) | 6/134 (4%) | 7/136 (5%) | 7/134 (5%) |
| Febrile | 0/136 (0%) | 1/134 (1%) | 0/136 (0%) | 1/134 (1%) |
| Gastro | 0/136 (0%) | 1/134 (1%) | 1/136 (1%) | 0/134 (0%) |
| Injury | 2/136 (1%) | 0/134 (0%) | 1/136 (1%) | 1/134 (1%) |
| Other | 3/136 (2%) | 2/134 (1%) | 2/136 (1%) | 3/134 (2%) |
| Other surgery | 1/136 (1%) | 0/134 (0%) | 0/136 (0%) | 1/134 (1%) |
| Respiratory | 0/136 (0%) | 2/134 (1%) | 0/136 (0%) | 2/134 (1%) |
| Skin / soft tissue infection | 3/136 (2%) | 4/134 (3%) | 3/136 (2%) | 4/134 (3%) |
| 12 months | Placebo | Cotrimoxazole | No Povidone-iodine | Povidone-iodine |
|  | N=138 | N=140 | N=138 | N=140 |
| ENT | 12/138 (9%) | 16/140 (11%) | 12/138 (9%) | 16/140 (11%) |
| Febrile | 1/138 (1%) | 1/140 (1%) | 1/138 (1%) | 1/140 (1%) |
| Gastro | 1/138 (1%) | 1/140 (1%) | 2/138 (1%) | 0/140 (0%) |
| Injury | 5/138 (4%) | 2/140 (1%) | 5/138 (4%) | 2/140 (1%) |
| Other | 8/138 (6%) | 4/140 (3%) | 5/138 (4%) | 7/140 (5%) |
| Other surgery | 0/138 (0%) | 4/140 (3%) | 2/138 (1%) | 2/140 (1%) |
| Respiratory | 3/138 (2%) | 3/140 (2%) | 1/138 (1%) | 5/140 (4%) |
| Skin / soft tissue infection | 1/138 (1%) | 3/140 (2%) | 4/138 (3%) | 0/140 (0%) |

**Table 8.** Audiology outcomes at 12 months (mean worse hearing ear in dBs)

|  | Povidone-iodine | No Povidone-iodine | Mean Diff | P-value | Co-trimoxazole | Placebo | Mean Diff | P-value |
| --- | --- | --- | --- | --- | --- | --- | --- | --- |
| Audiology/ear exam (%) | 65/120 (54%) | 68/118 (58%) |  |  | 62/120 (52%) | 71/118 (60%) |  |  |
| Mean worst ear in dB | 33.8 | 33.2 | -1.3 | 0.40 | 32.4 | 32.8 | -0.4 | 0.8 |
| SD | 10.3 | 9.4 | (-4.7, 2.1) |  | 9.4 | 9.4 | (-3.8, 3) |  |
| Adjusted# | 33.1 (6.05) | 31.6 (6.05) | 1.5 (-1.9, 4.9) | 0.38 | 32.9 (6.04) | 31.6 (6.04) | 1.3 (-2.1, 4.7) | 0.46 |

#Adjusted for baseline hearing outcomes

### Side effects

No children in the study developed an illness that required cessation of trial interventions or were withdrawn from the trial for adverse side effects. Mild side effects reported in 19/269 children included ‘upset stomach’ from the oral/placebo medication (10/136 active group; 9/134 placebo group). 7/133 children who had occasional, transient dizziness or ear discomfort, lasting up to 5 minutes following the povidone-iodine ear washes. Cessation of study treatment for children undergoing ear surgery was managed by the treating clinician.

### Audiology

An audiogram was performed at baseline in 168 (60%) children and at ∼12 months after randomisation in 133 (47%) children. 89 (32%) children had both baseline and 12-month audiograms. The mean hearing level in the worse hearing ear was around 33dB both before and after the intervention period (Tables 1,8). 3% of children had normal hearing in their worst ear before the intervention period. Comparisons of post-intervention hearing levels in the worse ear were: i) povidone-iodine 33dB (SD 10·1, n=111) versus no betadine 34dB (SD 9·8, n=107); mean difference −1 dB (95% confidence interval (CI) −4 to 3); and ii) cotrimoxazole 33dB (SD 9.7, n=108) versus placebo 33dB (SD 10·3, n=110); mean difference of −1 (95%CI −4 to 3) (Figures 4, 5). The unadjusted averages showed no statistical difference between treatment groups. Similar proportions of children with an audiologic assessment were clinical failures at 16 weeks (60%) when compared to the rest of the cohort. Mean hearing in the worst hearing ear was 33dB in children with and without discharge at 12 months.

### Microbiology

The microbiology of ear discharge across all groups (proportion of children positive) at baseline was anaerobic growth (23%), *P*. *aeruginosa* (21%), NTHi (17%) and *Proteus* spp. (16%). *S. aureus* (14%), fungi and yeast (12%), and *S. pneumoniae* (7%) were isolated. Presence of bacterial species of interest is reported by treatment group in Table 1. Post treatment at 16 weeks (n=265, including wet and dry ears), pneumococci (4%), NTHi (9%), *P. aeruginosa* (6%), *Proteus* species (7%) and any anaerobic growth (10%) decreased from baseline. Fungal or yeast growth increased (24%). *S. aureus* was stable (15%), as were coagulase negative staphylococci (27% to 33%). Culture negative swabs increased (3% to 6%). When including the wet ears only post-treatment (n=132), *P. aeruginosa* (11%), *Proteus* species (10%) and any anaerobic growth (18%) decreased from baseline, fungal or yeast growth increased (21%), pneumococci (7%), *S. aureus* (15%), NTHi (12%) and coagulase negative staphylococci (31%) were stable. *M. catarrhalis* was not isolated from ear discharge at either timepoint. There were no statistical differences in culture results between treatment groups (tables 4, 5; figures 2, 3; supplement 1, table 2) at 16 weeks.

## DISCUSSION

This is the first randomised controlled trial for treatment of CSOM using standard topical treatment plus povidone-iodine ear washes and oral antibiotics as adjunctive therapies.

In this RCT, oral cotrimoxazole increased the proportion of children with dry ears at 16 weeks, consistent with the outcomes of an earlier Dutch trial.^10^ Other oral interventions have not been similarly successful for CSOM. Oral amoxicillin^14,15^ in addition to topical treatment was used in two studies in African children with CSOM^14,15^. Minja *et al*^14^ reported increased proportions of children with dry ears in those receiving oral and topical boric acid treatment compared to controls (dry mopping alone) (32/57, 56% vs 34/110, 31%) at 3-4 months, however the oral amoxycillin had little additional impact in comparison to the topical boric acid group (56% vs 57/105, 54%). The second trial used oral amoxicillin and topical Sofradex.^15^ This combination was superior to dry mopping or no treatment alone for resolution of discharge (51% dry mopping plus topical antibiotics plus oral antibiotics vs 22% dry mopping or no treatment controls) at 16 weeks. This difference could be attributed to topical antibiotic treatment alone, therefore the benefit of the oral amoxycillin is not clear. Similarly, a trial comparing topical ciprofloxacin^16^ with combination topical and oral ciprofloxacin found no increased benefit of the oral treatment. These trials did not include microbiological assessments. The inclusion of oral cotrimoxazole has shown a clear benefit in this trial. The recent Cochrane review of systemic antibiotics for CSOM highlights that evidence in this area is limited and the authors make no strong recommendations.^17^ This study will add to the body of evidence for use of oral medication in addition to standard topical treatment.

### Clinical outcomes

This pragmatic trial reports increased cessation of discharge (16%) in children receiving up to 16 weeks of cotrimoxazole. We did not see a benefit (on average) at 16 weeks from adding povidone-iodine washes to standard topical treatment. However, the ear washes were well tolerated, and didn’t cause any worse outcomes. Whilst there was clinical improvement at 16 weeks, this benefit did not persist long-term and 57% of children had a wet perforation at the 12 month follow up. Additional efforts are required to increase positive outcomes for these children experiencing ongoing chronic ear discharge.

### Audiology

The Health Department outreach audiologists performed the baseline and 12 month follow-up hearing assessment. Due to the infrequency of their scheduled visits (once or twice a year per community) and their many competing responsibilities, less than 50% of scheduled audiology assessments were achieved. The substantial amount of missing data means that any estimates are at high risk of bias. It is also possible that the hearing tests were done at a time when the ear condition was different to the ear condition documented by their trial assessment. The lack of difference between overall hearing outcomes pre and post treatment could be that some perforations dried but had not healed (and that the ear discharge was not contributing much to the hearing loss). It is also possible that there was not much discharge present at the time of the baseline hearing test. The adjusted mean decibel hearing difference was small and was not statistically significant for either of the two main intervention comparisons (1.5dB for the povidone-iodine wash vs no wash, and 1.3dB for the antibiotic vs placebo group).

### Microbiology

Our previous randomised trial comparing topical ciprofloxacin with Sofradex for CSOM found that *Staphylococcus* species were the commonly isolated pathogens in ear discharge after treatment (present in 75% of specimens)^6^. This previous study also reported higher baseline proportions of *P*. *aeruginosa* (60%), and slightly higher *H. influenzae* (22%). Similarly, study from a similar population described higher baseline *P. aeruginosa* (48%) and respiratory pathogens were infrequently isolated (6/105 children).^8^ The change in *P. aeruginosa* isolation may be due to ciprofloxacin as the primary topical treatment. Microbiological environments also likely change with duration of discharge. Children had varying periods of discharge prior to enrolment (minimum of 6 weeks). Overall (compared to the previous trial), bilateral chronic discharge was less common and the perforation sizes were smaller.

Consistent with the Dutch trial, cotrimoxazole did not impact the isolation of *H. influenzae*, *S. pneumoniae*, or other otopathogens from ear discharge in this study.^10^ There was a non-significant difference in NTHi in the povidone-iodine group, compared to the no povidone-iodine group (11% vs 6%). Overall, proportions of otopathogens and other OM associated organisms were lower post treatment at 16 weeks. This reduction could be due to the increased proportion of dry ears, making canal flora more commonly isolated than pathogens. Inclusion in this trial could have also increased management of CSOM, such as ear toilet (tissue spears) by families, and more frequent assessment (contact with research staff during the intervention period and primary healthcare for additional study drugs and ciprofloxacin), assisting discharge cessation through increased management.

Several observational studies of CSOM associated microbiology have been published recently. Overall, they report culture of varying predominant organisms, including *S. aureus, P*. *aeruginosa,* fungal species, and proteus species. ^18–26^ These studies all used aerobic culture methods. Culturing for retrieval of anaerobic species or fungi / yeasts was not consistent. Bacterial profiles associated with CSOM are predominantly polymicrobial with a range of micro-organisms isolated from ear discharge.

## CONCLUSION

Oral cotrimoxazole in addition to standard topical treatment for up to 16 weeks increased cessation of discharge compared to placebo. Povidone-iodine ear wash in addition to standard therapy was not effective in our study. Our analysis of children at ∼12 months follow-up shows the very high level of persistent ear discharge and hearing loss in children with CSOM. It appears that the addition of oral cotrimoxazole to standard topical treatment can reduce the time with CSOM while they are receiving oral antibiotics. However, these children remain at risk of further ear discharge and CSOM in the future. In the long-term (12 months after randomisation) around 60% of children still had CSOM. The adjunctive interventions we assessed in this trial did not appear to have much impact on this outcome. In some populations, CSOM remains very difficult to treat effectively and the burden of disease over a lifetime remains substantial. While prevention of this condition is most important, further trials of different combinations of available treatments and alternative approaches to management are needed.

## Trial Status

Complete. Recruitment commenced in February 2015 and finished in December 2018.

## List of abbreviations

AOMwiP: Acute otitis media with perforation
AOMwoP: Acute otitis media without perforation
CSOM: Chronic suppurative otitis media
DP: Dry perforation
GCP: Good Clinical Practice
ITT: Intention-to-treat
IPD: Invasive pneumococcal disease
NHMRC: National Health and Medical Research Council
NT: Northern Territory
NTHi: Non-typeable Haemophilus influenzae
OM: Otitis media
OME: Otitis media with effusion
TMP: Tympanic membrane perforation

## Declarations

### Ethics approval and consent to participate

Approval by the Human Research Ethics Committee of the Northern Territory Department of Health and the Menzies School of Health Research (HREC-2014-2170), and the Central Australian Human Research Ethics Committee (CAHREC-14-228) was granted to conduct this study in the NT. Each participating community also provided approval of the study to be conducted and informed written consent was obtained from every subject’s parent/guardian prior to enrolment.

Trial registration: ACTRN12614000234617 at ANZCTR, registered April 2014

### Consent to publish

There are no individual persons’ data of any form that has been included in this manuscript.

### Availability of data and materials

Data that supports this study protocol can be obtained from the corresponding author, providing appropriate ethical and community approvals are provided. Data are not available on a public repository.

### Authors’ contributions

PM conceptualised the study design, acquired the funding and is accountable for the accuracy and integrity of this body of work. AJL made substantial contributions to the funding application, study concept and design. JB established and maintained all database and laboratory components of the study. JB, VO, PSM and CW drafted and prepared the manuscript. CW is responsible for all other aspects of the clinical trial including accurate acquisition and maintenance of the trial data. VO and MC contributed to the design of the data analysis plan. VO did the analysis with assistance from JB. RL, SN, HP, KC, HC, KE, PT and ST contributed to the clinical design aspects of the study. HSV and JB designed the laboratory analysis plan. All authors read and approved the final manuscript.

## Acknowledgements

We thank all the families and children who participated in this study, the health service staff and community members who assisted with the research, as well as the clinical, laboratory and community researchers for driving the project. We appreciate the support of the NT urban and remote Indigenous councils and traditional owners for allowing us to conduct this study in their communities.

