## Supplemental tables and method for "Povidone-iodine ear wash and oral cotrimoxazole for chronic suppurative otitis media in Australian Aboriginal children: a randomised controlled 2x2 factorial design trial"

**Table 1:** Baseline characteristics by randomisation group

| | | Povidone-iodine | No Povidone-iodine | Co-trimoxazole | Placebo |
|---|---|---|---|---|---|
| | | n=140 | n=139 | n=140 | n=139 |
| Gender | Male | 66/140 (47%) | 69/139 (50%) | 75/140 (54%) | 60/139 (43%) |
| | Female | 74/140 (53%) | 70/139 (50%) | 65/140 (46%) | 79/139 (57%) |
| Age (mean, years (SD)) | | 7 (4) | 7 (4) | 8 (4) | 7 (4) |
| Age (year group) | <4yrs | 40/140 (29%) | 30/139 (22%) | 36/140 (26%) | 34/139 (24%) |
| | 4-10yrs | 69/140 (49%) | 69/139 (50%) | 72/140 (51%) | 66/139 (47%) |
| | >10yrs | 31/140 (22%) | 40/139 (29%) | 32/140 (23%) | 39/139 (28%) |
| Weight (kg) (SD) | | 23 (13) | 25 (14) | 23 (14) | 24 (13) |
| CSOM category | Unilateral | 95/140 (68%) | 89/139 (64%) | 90/140 (64%) | 94/139 (68%) |
| | Bilateral | 45/140 (32%) | 50/139 (36%) | 50/140 (36%) | 45/139 (32%) |
| Worst perforation | 3-10% | 40/140 (29%) | 35/139 (25%) | 43/140 (31%) | 32/139 (23%) |
| | 11-25% | 47/140 (34%) | 48/139 (35%) | 44/140 (31%) | 51/139 (37%) |
| | 26-50% | 36/140 (26%) | 34/139 (24%) | 34/140 (24%) | 36/139 (26%) |
| | 51-75% | 11/140 ( 8%) | 15/139 (11%) | 13/140 ( 9%) | 13/139 ( 9%) |
| | >75% | 6/140 ( 4%) | 7/139 ( 5%) | 6/140 ( 4%) | 7/139 ( 5%) |
| Median worst perforation size % (IQR) | | 20 (8-35) | 20 (8-35) | 22 (10-38) | 20 (8-35) |
| Hearing (worst) | | n=79 | n=89 | n=85 | n=83 |
| normal | | 12/79 (15%) | 8/89 ( 9%) | 10/85 (12%) | 10/83 (12%) |
| mild | | 26/79 (33%) | 28/89 (31%) | 29/85 (34%) | 25/83 (30%) |
| moderate | | 39/79 (49%) | 53/89 (60%) | 45/85 (53%) | 47/83 (57%) |
| severe | | 2/79 ( 3%) | 0/89 ( 0%) | 1/85 ( 1%) | 1/83 ( 1%) |
| Mean worst hearing loss dB (SD) | | 32 (11) | 34 (10) | 33 (11) | 33 (11) |
| Ear discharge | | n=139 | n=137 | n=139 | n=137 |
| *P. aeruginosa* | | 31/139 (22%) | 28/137 (20%) | 24/139 (17%) | 35/137 (26%) |
| NTHi | | 19/139 (14%) | 27/137 (20%) | 21/139 (15%) | 25/137 (18%) |
| Swarming proteus | | 24/139 (17%) | 20/137 (15%) | 19/139 (14%) | 25/137 (18%) |
| *S. aureus* | | 12/139 ( 9%) | 27/137 (20%) | 20/139 (14%) | 19/137 (14%) |
| *S. pneumoniae* | | 8/139 ( 6%) | 10/137 ( 7%) | 10/139 ( 7%) | 8/137 ( 6%) |
| Anaerobic growth | | 30/139 (22%) | 34/137 (25%) | 41/139 (29%) | 23/137 (17%) |
| Fungi or yeast | | 15/139 (11%) | 18/137 (13%) | 17/139 (12%) | 16/137 (12%) |

| | | | | |
|---|---|---|---|---|
| Coag-neg Staph | 38/139 (27%) | 37/137 (27%) | 38/139 (27%) | 37/137 (27%) |
| Any growth* | 132/139 (95%) | 137/137 (100%) | 135/139 (97%) | 134/137 (98%) |

*Any growth is defined as any cultured bacteria or fungi/yeasts;

**Table 2: Clinical and microbiologic outcomes by randomised group at 16 weeks, including primary outcomes (no hearing outcomes assessed at 16 weeks)**

| 16 weeks | povidone-iodine | No povidone-iodine | p-value | Cotrimoxazole | Placebo | p-value |
|---|---|---|---|---|---|---|
| | N=134 | N=136 | | N=134 | N=136 | |
| Clinical failure | | | | | | |
| No | 68/134 (51%) | 67/136 (49%) | 0·81 | 78/134 (58%) | 57/136 (42%) | 0·007 |
| Yes | 66/134 (49%) | 69/136 (51%) | | 56/134 (42%) | 79/136 (58%) | |
| Worst ear | | | | | | |
| Normal | 2/134 ( 1%) | 4/136 ( 3%) | 0·63 | 4/134 ( 3%) | 2/136 ( 1%) | 0·080 |
| OME | 13/134 (10%) | 7/136 ( 5%) | | 10/134 ( 7%) | 10/136 ( 7%) | |
| AOMwiP | 2/134 ( 1%) | 2/136 ( 1%) | | 1/134 ( 1%) | 3/136 ( 2%) | |
| Dry perf | 53/134 (40%) | 56/136 (41%) | | 64/134 (48%) | 45/136 (33%) | |
| CSOM | 64/134 (48%) | 67/136 (49%) | | 55/134 (41%) | 76/136 (56%) | |
| CSOM | | | | | | |
| Unilateral | 40/64 (63%) | 48/67 (72%) | 0·27 | 36/55 (65%) | 52/76 (68%) | 0·72 |
| Bilateral | 24/64 (38%) | 19/67 (28%) | | 19/55 (35%) | 24/76 (32%) | |
| Worst perforation | | | | | | |
| Intact | 18/132 (14%) | 13/135 (10%) | 0·45 | 15/132 (11%) | 16/135 (12%) | 0·76 |
| <=2% | 3/132 ( 2%) | 3/135 ( 2%) | | 2/132 ( 2%) | 4/135 ( 3%) | |
| 3-10% | 36/132 (27%) | 27/135 (20%) | | 35/132 (27%) | 28/135 (21%) | |
| 11-25% | 31/132 (23%) | 38/135 (28%) | | 34/132 (26%) | 35/135 (26%) | |
| 26-50% | 25/132 (19%) | 37/135 (27%) | | 32/132 (24%) | 30/135 (22%) | |
| 51-75% | 15/132 (11%) | 15/135 (11%) | | 12/132 ( 9%) | 18/135 (13%) | |
| >75% | 4/132 ( 3%) | 2/135 ( 1%) | | 2/132 ( 2%) | 4/135 ( 3%) | |
| Ear discharge | | | | | | |
| *P. aeruginosa* | 10/134 ( 7%) | 6/131 ( 5%) | 0·32 | 9/129 ( 7%) | 7/136 ( 5%) | 0·53 |
| NTHi | 15/134 (11%) | 8/131 ( 6%) | 0·14 | 12/129 ( 9%) | 11/136 ( 8%) | 0·73 |
| *Proteus* | 8/134 ( 6%) | 10/131 ( 8%) | 0·59 | 8/129 ( 6%) | 10/136 ( 7%) | 0·71 |
| *S. aureus* | 18/134 (13%) | 21/131 (16%) | 0·55 | 19/129 (15%) | 20/136 (15%) | 1·00 |
| Coag-neg Staph | 51/134 (38%) | 38/131 (29%) | 0·12 | 48/129 (37%) | 41/136 (30%) | 0·22 |
| Fungi or yeast | 34/134 (25%) | 31/131 (24%) | 0·75 | 36/129 (28%) | 29/136 (21%) | 0·21 |
| *S. pneumoniae* | 6/134 ( 4%) | 4/131 ( 3%) | 0·54 | 7/129 ( 5%) | 3/136 ( 2%) | 0·17 |
| No growth | 7/134 ( 5%) | 10/131 ( 8%) | 0·42 | 9/129 ( 7%) | 8/136 ( 6%) | 0·72 |
| Anaerobic growth | 15/134 (11%) | 13/131 (10%) | 0·74 | 13/129 (10%) | 15/136 (11%) | 0·80 |

**Table 3: Clinical and hearing outcomes by randomised group at 12 months, including primary outcomes (no microbiologic outcomes available for 12 months)**

| 12 months | Betadine | No betadine | p-value | Cotrimoxazole | Placebo | p-value |
|---|---|---|---|---|---|---|
| | N=140 | N=138 | | N=140 | N=138 | |
| Clinical failure | | | | | | |
| No | 54/140 (39%) | 66/138 (48%) | 0·12 | 58/140 (41%) | 62/138 (45%) | 0·56 |
| Yes | 86/140 (61%) | 72/138 (52%) | | 82/140 (59%) | 76/138 (55%) | |
| Worst ear | | | | | | |
| normal | 3/139 ( 2%) | 4/132 ( 3%) | 0·24 | 5/136 ( 4%) | 2/135 ( 1%) | 0·66 |
| OME | 15/139 (11%) | 12/132 ( 9%) | | 14/136 (10%) | 13/135 (10%) | |
| AOMwiP | 4/139 ( 3%) | 2/132 ( 2%) | | 4/136 ( 3%) | 2/135 ( 2%) | |
| Dry perf | 32/139 (23%) | 44/132 (33%) | | 34/136 (25%) | 42/135 (31%) | |
| AOMwoP | 3/139 ( 2%) | 0/132 ( 0%) | | 1/136 ( 1%) | 2/135 ( 2%) | |
| CSOM | 82/139 (59%) | 70/132 (53%) | | 78/136 (57%) | 74/135 (55%) | |
| Unilateral | 55/82 (67%) | 46/70 (66%) | 0·86 | 48/78 (62%) | 53/74 (72%) | 0·19 |
| Bilateral | 27/82 (33%) | 24/70 (34%) | | 30/78 (38%) | 21/74 (28%) | |
| Worst perforation | | | | | | |
| intact | 27/138 (20%) | 17/128 (13%) | 0·25 | 22/134 (16%) | 22/132 (17%) | 0·74 |
| <=2% | 3/138 ( 2%) | 3/128 ( 2%) | | 4/134 ( 3%) | 2/132 ( 2%) | |
| 3-10% | 27/138 (20%) | 19/128 (15%) | | 23/134 (17%) | 23/132 (17%) | |
| 11-25% | 33/138 (24%) | 40/128 (31%) | | 39/134 (29%) | 34/132 (26%) | |
| 26-50% | 37/138 (27%) | 30/128 (23%) | | 30/134 (22%) | 37/132 (28%) | |
| 51-75% | 6/138 ( 4%) | 14/128 (11%) | | 9/134 ( 7%) | 11/132 ( 8%) | |
| >75% | 5/138 ( 4%) | 5/128 ( 4%) | | 7/134 ( 5%) | 3/132 ( 2%) | |
| Worst hear | | | | | | |
| Normal | 5/68 ( 7%) | 9/66 (14%) | 0·40 | 9/71 (13%) | 5/63 ( 8%) | 0·39 |
| Mild | 28/68 (41%) | 22/66 (33%) | | 23/71 (32%) | 27/63 (43%) | |
| Moderate | 35/68 (51%) | 35/66 (53%) | | 39/71 (55%) | 31/63 (49%) | |

**Table 4: Clinical failure at 12 months in povidone-iodine and cotrimoxazole group**

| Clinical failure | Betadine | No betadine | RR | RD | p |
|---|---|---|---|---|---|
| Unadjusted | 86/140 (61%) | 72/138 (52%) | 1·18(0·96,1·45) | 9 (-2,21) | 0·12 |
| Adjusted | 86/140 (61%) | 72/138 (52%) | 1·17(0·95,1·44) | 8 (-3,20) | 0·13 |
| | Antibiotics | Placebo | | | |
| Unadjusted | 82/140 (59%) | 76/138 (55%) | 1·06(0·87,1·31) | 4(-8,15) | 0·56 |
| Adjusted | 82/140 (59%) | 76/138 (55%) | 1·09(0·89,1·33) | -16(-28,-4) | 0·41 |

**Table 5:** Serious Adverse Events summary by randomisation group

| SAE type | Pov / placebo | | Pov / cotrimoxazole | | No Pov / placebo | | No Pov / cotrimoxazole | | Total =69 |
|---|---|---|---|---|---|---|---|---|---|
| Intervention stage | <16wks | >16wks | <16wks | >16wks | <16wks | >16wks | <16wks | >16wks | |
| ENT | 6 | 7 | 2 | 6 | 1 | 3 | 3 | 4 | 32 |
| Fever | 0 | 0 | 2 | 0 | 0 | 1 | 0 | 0 | 3 |
| Gastrointestinal | 0 | 0 | 0 | 0 | 0 | 0 | 1 | 1 | 2 |
| Injury | 0 | 2 | 0 | 0 | 1 | 0 | 0 | 0 | 3 |
| Other | 0 | 1 | 1 | 2 | 0 | 3 | 0 | 0 | 7 |
| Other surgery | 1 | 0 | 0 | 2 | 0 | 0 | 0 | 2 | 5 |
| Respiratory | 1 | 4 | 2 | 2 | 0 | 0 | 0 | 1 | 10 |
| Skin / tissue infection | 2 | 0 | 0 | 0 | 1 | 1 | 1 | 2 | 7 |

Table 6. Dominant species in ear discharge of worst CSOM ear at baseline and worst discharging ear (CSOM or AOM) at 16 weeks

| Dominant organism- Baseline | | | Dominant organism- 16 weeks | | |
|---|---|---|---|---|---|
| | n | % | | n | % |
| Corynebacterium spp | 101 | 36 | Corynebacterium spp | 51 | 39 |
| Diphtheroid spp | 50 | 18 | Other | 21 | 16 |
| Pseudomonas spp | 35 | 13 | Pseudomonas spp | 15 | 11 |
| Proteus spp | 31 | 11 | Diphtheroid spp | 13 | 10 |
| Other | 16 | 6 | Proteus spp | 10 | 8 |
| Coliform | 10 | 4 | Coliform | 5 | 4 |
| *S. aureus* | 9 | 3 | No growth | 2 | 2 |
| No growth | 8 | 3 | *H. influenzae* | 4 | 3 |
| Coagulase negative *Staphylococcus* | 5 | 2 | Coagulase negative *Staphylococcus* | 4 | 3 |
| *H. influenzae* | 4 | 1 | *S. aureus* | 3 | 2 |
| βHS | 3 | 1 | Fungi / yeast | 2 | 2 |
| Fungi / yeast | 3 | 1 | βHS | 1 | 1 |
| *S. pneumoniae* | 2 | 1 | *S. pneumoniae* | 0 | 0 |
| αHS not Spn | 2 | 1 | αHS not Spn | 0 | 0 |
| Total | 279 | 100 | Total | 132 | 100 |

HS=haemolytic streptococci, Spn= *S. pneumoniae*

Table 7: Dominant species in worst ear discharge at 16 weeks by randomisation groups

| Dominant organism | Pov / placebo n=40 | Pov / cotrimoxazole n=25 | No Pov / placebo n=39 | No Pov / cotrimoxazole n=27 | Total n=133 |
|---|---|---|---|---|---|
| Corynebacterium spp | 19 | 6 | 13 | 13 | 51 |
| Other | 9 | 5 | 5 | 2 | 21 |
| Pseudomonas spp | 4 | 4 | 4 | 3 | 15 |
| Diphtheroid spp | 1 | 3 | 5 | 4 | 13 |
| Proteus spp | 2 | 3 | 5 | 0 | 10 |
| Coliform | 1 | 0 | 2 | 2 | 5 |
| No growth | 0 | 3 | 0 | 1 | 4 |
| *H. influenzae* | 2 | 0 | 1 | 1 | 4 |
| Coagulase negative *Staphylococcus* | 1 | 1 | 2 | 0 | 4 |
| *S. aureus* | 1 | 0 | 1 | 1 | 3 |
| Fungi / yeast | 0 | 1 | 0 | 1 | 2 |
| βHS | 0 | 0 | 1 | 0 | 1 |

**Microbiological methods**

Capsular Spn were identified by morphology, optochin resistance and a positive Quellung reaction (Statens Serum Institute of Copenhagen, Denmark). Presumptive NTHi were identified by morphology, X and V dependence (Oxoid) and coagglutination negative (Haemophilus Phadebact, Remel). Mc were identified by morphology, oxidase production and Gram stain. Sa were identified by colony morphology and coagulation by latex agglutination (Staphaurex, Remel). βeta-lactamase production by NTHi and Mc was determined using nitrocephin (Oxoid, Australia). Pa were identified by oxidase production, growth on pseudomonas agar at 42$^{o}$C, and API 20NE (BioMerieux). Antimicrobial susceptibility determination was done by the EUCAST method. Susceptibility was determined for Spn (oxacillin, tetracycline, erythromycin, sulphamethoxazole trimethoprim, chloramphenicol, and ciprofloxacin), NTHi (ampicillin, sulphamethoxazole trimethoprim, ciprofloxacin, and erythromycin), Sa (oxacillin, penicillin, erythromycin, ciprofloxacin, gentamycin, sulphamethoxazole trimethoprim, and cefoxitin) and Pa (ciprofloxacin, gentamycin, ceftazidime, sulphamethoxazole trimethoprim, piperacillin/tazobactam and meropenem).
